# Cannabis use in relation to Pulse Pressure and Mean Arterial Pressure in US Adults

**DOI:** 10.1101/2023.05.12.23289921

**Authors:** Stella Ruddy, Vincenzo Di Marzo, Gerard Ngueta

**Affiliations:** Centre de recherche du CHU de Sherbrooke, Division d’endocrinologie, Québec (Canada); Chaire d’excellence en recherche du Canada sur l’axe microbiome-endocannabinoïdome en santé métabolique (CERC-MEND), Centre de recherche de l’Institut de cardiologie et de pneumologie de Québec, Faculté de médecine, Département de médecine, Université Laval, Québec, Canada; Unité Mixte Internationale en Recherche Chimique et Biomoléculaire sur le Microbiome et son Impact Sur la Santé Métabolique et la Nutrition (UMI-MicroMeNu), Université Laval and Consiglio Nazionale delle Ricerche, Istituto di Chimica Biomolecolare, Pozzuoli, Italy; Université de Sherbrooke, Faculté de médecine, Département des sciences de la santé communautaire

## Abstract

**Background:** Pulse pressure (PP) and mean arterial pressure (MAP) have been well-established as markers of cardiovascular risk in clinical settings. We aimed to determine the impact of cannabis use on both PP and MAP in U.S. adults and to assess the modifying role of sex.

**Methods:** We abstracted data from the 2009 to 2018 National Health and Nutrition Examination survey (NHANES). Cannabis use was assessed by NHANES professionals in adults aged 18 to 59 years by using computer-assisted self-interviews. We defined PP as the difference between systolic and diastolic BP, and MAP as diastolic BP plus one third of PP. We used multivariable linear models to estimate the covariates-adjusted associations and assessed effect modification by including sex×exposure interaction terms into the model.

**Results:** The mean age of the study population (n=8,942) was 35.0±11.9 years, with 51% female (n=4,551). Mean±SD PP and MAP were 46±13 mm Hg and 82±13 mm Hg, respectively. We found a significant interaction between sex and cannabis use in relation to PP (*P*=0.0878) and no interaction when modeling MAP (*P*=0.2084). The mean difference of PP between cannabis users and never-users increased with the frequency of use per week, being +4.5 mm Hg (*P*=0.0004) in those who reported 1 use/week, +4.9 mm Hg (*P*<0.0001) for 2-3 uses/week and +4.9 mm Hg (*P*<0.0001) for ≥ 4 uses/week. In females, only those who reported ≥ 4 uses/week showed a higher PP (+3.1 mm Hg; *P*=0.0050) compared with never-users.

**Conclusions:** In US adults aged 18 to 59 years, cannabis use is associated with widening of PP in males.

**CLINICAL PERSPECTIVES:** *What is new?:* We first investigated the cannabis use in relation to PP and MAP and found that cannabis use is associated with widened PP in sex-specific manner.

*What are the clinical implications?:* Further evidence from cohort studies is required before it can be firmly concluded that cannabis use is linked to increased PP. Patients should stop cannabis use to optimize treatments with reduction of PP as specific therapeutic target.

---

Over the last decades, cardiovascular diseases (CVD) have been the leading cause of death in high income countries.^1^ Elevated blood pressure (BP) is a major and modifiable risk factor for CVD, and evidence suggest that wide pulse pressure (PP) is a precursor of isolated systolic hypertension and early predictor of incident CVD.^2^ In previous investigations, PP and mean arterial pressure (MAP) has been well-established as markers of CVD risk in clinical settings^3^ and predictors of mortality.^4^

As of early 2022, almost 20 US states legalized the recreational use of cannabis for adults. The timeline of decriminalizing cannabis use coincides with the aging of the baby boomer’s generation, with monotonic increase rates of cannabis use among ageing US adults.^5^ Cannabis is now listed in the Top 3 of the most used psychoactive substances in the United States, with over 48.2 million people aged 12 or older reporting past-year cannabis use (vs. 25.8 million in 2002).^6^ According to a 2023 study, cannabis-related emergency department visits are increasing among older adults in California,^7^ pointing to the cannabis study as a priority research area on the public health agenda.

The cannabis plant contains hundreds different compounds, including cannabinoids.^8^ The most known include delta-tetrahydrocannabinol (THC), cannabinol (CBN), and cannabidiol (CBD). THC is responsible for the psychoactive properties of cannabis,^9^ while CBD presents vasorelaxation actions in arteries.^10^ THC exerts its action through at least two G-protein-coupled cannabinoid receptors (CB1 and CB2). CB1 was first identified in 1990 as a target of THC,^11^ and CB2 in 1993.^12^ Evidence suggest that CB1 and CB2 receptors may modulate cardiometabolic risk and atherogenesis and may protect cardiomyocytes from damage.^13^ In rats, acute administration of THC and endogenous agonists of CB1 receptors results in bradycardia and lowering of arterial pressure through CB1 in a dose-dependent manner.^14 15^ CB2 receptors are expressed in cardiomyocytes, coronary endothelial cells and smooth muscle cells, and are suggested to play a cardioprotective role.^16^ It can be hypothesized that the risk of elevated PP can be reduced in cannabis users.

Results from cross-sectional studies suggest a high prevalence of elevated BP in cannabis users, as compared with never users.^17^ However, this finding is not consistent across studies, and modest^18^ or inverse associations have been reported elsewhere.^19^ In a prospective study of hypertensive patients, Abuhasira et al.^20^ found that cannabis therapy for 3 months was associated with a reduction in 24-hours systolic and diastolic BP. A randomized, double-blind, crossover study including nine healthy male volunteers suggests that acute administration of CBD reduces resting systolic BP and stroke volume, with increased heart rate and maintained cardiac output.^21^ Abrupt cessation of heavy cannabis use has been associated with clinically significant increase in both systolic and diastolic BP,^22^ but another study found no changes after quitting.^23^ As a whole, results remain inconsistent across studies, independent of the study design. This might be due to different composition in THC, CBD and other cannabinoids of the various cannabis variants used by cannabis users.

Previous studies suggest sex-related differences in cardiovascular response to cannabis exposure.^24^ The amount of THC reaching the blood after smoking also differ between male and female.^25^ Evidence from animal models suggest that gonadal hormones influence cannabinoid receptor density in a sex-dependent manner.^26^ In our knowledge, the potential association of cannabis exposure with PP and MAP has not been investigated, and sex-differences in this relationship remain unknown. To address this gap of knowledge, here, we conducted cross-sectional analyses to determine the impact of cannabis use on both PP and MAP in U.S. adults and assessed the modifying role of sex.

## METHODS

### Study design and population study

We abstracted data from the 2009 to 2018 National Health and Nutrition Examination survey (NHANES),^27^ an ongoing cross-sectional survey of non-institutionalized US civilian participants. This nationally representative survey is conducted year after year by the National Center for Health Statistics (NCHS) and supported by the US Centers for Disease Control and Prevention (U.S. CDC). The data related to cannabis use were available for participants 18 to 59 years of age. All participants provided written informed consent and the NCHS Research Ethics Review Board approved all protocols. In this study, we excluded pregnant individuals (n=775), those under antihypertensive medication (n=5,612), those with known diabetes (n=998) or hypertriglyceridemia (n=1,325) and those with known CVD (n=271). We further excluded those who did not provide data on cannabis use (n=5,560). After all these exclusions, 8,942 individuals were included in the analysis.

### Medical evaluation and definitions

BP was measured by trained physicians using a standardized medical protocol.^28^ After 5 minutes of seated rest, three brachial BP measurements were obtained at 30-second intervals using a mercury sphygmomanometer (Bauman true gravity mercury wall model) with appropriate cuff sizes. For 2017-2018, the examiner measured BP using a validated oscillometric device (Omron 907 XL). The mean of the three was used to define systolic and diastolic BP for each participant. We defined PP as the difference between systolic and diastolic BP, and MAP as diastolic BP plus one third of PP.

### Assessing cannabis exposure

The NHANES questionnaire was administrated by the NHANES professionals during the household interview. To identify “Never users”, we referred to the question “Have you ever, even once, used marijuana or hashish?” (Yes/No). In those who reported current or past cannabis exposure, we next referred to the question “During the time that you smoked marijuana or hashish, how often would you usually use it?” to estimate the frequency of exposure (number of uses per week).

### Covariates

Covariates were selected a priori based on availability of variables and potential confounding that may affect both cannabis use and blood pressure measures. We included age at interview (continuous), sex (male vs. female), self-report race/ethnicity (non-Hispanic white, non-Hispanic black, Hispanic, Other), education level (less than high school, high school, some college/university), poverty-to-income ratio (continuous), acculturation score (continuous), serum cotinine (continuous), physical activity (hours/week) and NHANES cycles. The acculturation score was constructed based on three proxy measures: country of birth, length of time in the US and language spoken at home.^29^ For physical activity, we combined self-reported recreational and work-related moderate-to-vigorous activities. To avoid underestimation bias of our estimators, we did not further adjust for body mass index due to recent studies suggesting cannabis use and/or cannabis metabolites as predictors of body mass index in humans^30-32^ and animal models.^33^

### Statistical analysis

We conducted descriptive analyses in the overall sample and by exposure status to summarize the characteristics of the participants. We presented results from descriptive analyses as number (percentage) for discrete variables and as mean (±SD) or median (interquartile range [IQR]) for continuous variables. We used analysis of covariances (ANCOVA) models to describe means and 95% confidence intervals (95% CI) of both PP and MAP across exposure groups. We further categorized current users by frequency of use (1 use/week, 2-3 uses/week, and ≥ 4 uses/week) and cut-off points for frequencies of use were chosen to ensure both sufficient size per stratum and parsimonious regression models. We used multivariable linear models to estimate the covariates-adjusted associations between cannabis exposure groups and outcomes. To assess effect modification by sex, we further included the sex×exposure interaction term into the model. In case of significant multiplicative interaction, we performed stratified analyses and showed results by sex.

The unequal probability of being recruited and for non-response is common to surveys with complex designs, including NHANES.^34^ To deal with potential selection bias, the U.S. CDC developed weight variables,^35^ to account for the unequal probability of sampling and non-response. We used interview weight years and weights were adjusted for the inclusion of multiple surveys, by dividing the weight variable by the number of cycles used in our analyses. All analyses were conducted using SAS 9.4 (SAS Institute, Inc.), and *P* < 0.05 was considered for statistical significance. We considered interaction terms as statistically significant at the 0.10 probability level.

## RESULTS

The mean age of the study population (n=8,942) was 35.0±11.9 years, with 51% female (n=4,551) and 34% non-Hispanic white (n=3,062). Half of them earned a college or university degree (51%, 4,553) and current tobacco smokers represented 28% of the sample (n=2,518). Mean±SD systolic BP, diastolic BP, PP and MAP were 112±17 mm Hg, 67±13 mm Hg, 46±13 mm Hg and 82±13 mm Hg, respectively. Participants’ characteristics by cannabis exposure groups are shown in **Table 1**. In general, never users tended to have lower PP (44.3±12.4 mm Hg) and were less physically active (median of 40 hours/week).

**Table 1:** Characteristics of participants

|  | Never users | Former users | Current users |  |  |
| --- | --- | --- | --- | --- | --- |
|  |  |  | 1 use/week | 2-3 uses/week | ≥ 4 uses/week |
| **No. of participants** | 5,901 | 1,702 | 399 | 323 | 617 |
| **Age, years** | 35.5±11.9 | 36.2±12.0 | 31.2±11.9 | 30.5±10.4 | 31.2±10.7 |
| **Sex (n, % female)** | 3,357 (56.9) | 692 (40.7) | 174 (43.6) | 108 (33.4) | 220 (35.7) |
| **Race/Ethnicity** |  |  |  |  |  |
| Non-Hispanic white | 1,642 (27.8) | 855 (50.2) | 133 (33.3) | 149 (46.1) | 283 (45.9) |
| Non-Hispanic black | 1,028 (17.4) | 392 (23.0) | 135 (33.8) | 83 (25.7) | 183 (29.7) |
| Hispanic | 2,018 (34.2) | 299 (17.6) | 89 (22.3) | 60 (18.6) | 96 (15.5) |
| Other | 1,213 (20.6) | 156 (9.2) | 42 (10.6) | 31 (9.6) | 55 (8.9) |
| **Education level** |  |  |  |  |  |
| Less than high school | 1,114 (21.4) | 234 (14.6) | 77 (23.3) | 60 (21.5) | 119 (21.6) |
| High school | 1,026 (19.7) | 441 (27.6) | 105 (31.8) | 90 (32.1) | 157 (28.6) |
| Some college/university | 3,076 (58.9) | 925 (57.8) | 148 (44.9) | 130 (46.4) | 274 (49.8) |
| Missing values | 685 | 102 | 69 | 43 | 67 |
| **Poverty**<sup>*</sup> |  |  |  |  |  |
| No | 3,960 (75.1) | 1,218 (75.8) | 224 (62.8) | 201 (66.6) | 386 (69.3) |
| Yes | 1,314 (24.9) | 388 (24.2) | 133 (37.2) | 101 (33.4) | 171 (30.7) |
| Missing values | 627 | 96 | 42 | 21 | 60 |
| **Acculturation level** |  |  |  |  |  |
| Low | 2,122 (36.0) | 74 (4.3) | 32 (8.0) | 22 (6.8) | 19 (3.1) |
| Moderate | 1,898 (32.2) | 688 (40.4) | 168 (42.1) | 153 (47.4) | 318 (51.5) |
| High | 1,878 (31.8) | 940 (55.2) | 199 (49.9) | 148 (45.8) | 280 (45.4) |
| Missing values | 3 | 0 | 0 | 0 | 0 |
| **Current tobacco smokers** |  |  |  |  |  |
| No | 5,135 (87.0) | 848 (49.8) | 150 (37.6) | 115 (35.6) | 176 (28.5) |
| Yes | 766 (13.0) | 854 (50.2) | 249 (62.4) | 208 (64.4) | 441 (71.5) |
| **Physical activity, hours/week**<sup>†</sup> | 40.0 (16.0-104.0) | 61.0 (24.0-160.0) | 64.0 (26.0-176.0) | 84.0 (36.0-176.0) | 80.0 (24.0-224.0) |
| **Pulse pressure, mm Hg**<sup>‡</sup> | 44.3±12.4 | 45.3±12.5 | 46.8±12.4 | 49.0±13.3 | 47.9±14.1 |
| **Mean arterial pressure, mm Hg**<sup>§</sup> | 81.9±12.3 | 82.5±12.9 | 81.4±11.4 | 82.1±12.0 | 81.5±12.8 |

<sup>\*</sup> Defined as a poverty-to-income ratio less than 1
<sup>†</sup> Median (interquartile range)
<sup>‡</sup> Defined as difference between systolic and diastolic blood pressure
<sup>§</sup> Defined as diastolic blood pressure plus one third of pulse pressure

Association models are shown in **Table 2**. Compared with never users, the mean PP was significantly higher in current smokers and adjusted mean differences increased with frequency of use (+2.8 mm Hg, +3.5 mm Hg and +3.7 mm Hg for those reporting 1 use/week, 2-3 uses/week and ≥ 4 uses/week, respectively). There was no evidence of association between cannabis use and MAP.

**Table 2:** Adjusted regression coefficients (95% confidence intervals) for the associations of cannabis use with pulse pressure (PP) and mean arterial pressure (MAP) in US adults, NHANES 2009-2018

|  | PP |  |  |  | MAP |  |  |  |
| --- | --- | --- | --- | --- | --- | --- | --- | --- |
|  | Model 1<sup>*</sup> | P | Model 2<sup>†</sup> | P | Model 1<sup>*</sup> | P | Model 2<sup>†</sup> | P |
| Never users | Reference |  | Reference |  | Reference |  | Reference |  |
| Former users | -0.19 (-0.90, 0.51) | 0.5886 | 0.11 (-0.78, 1.00) | 0.8130 | 0.43 (-0.23, 1.09) | 0.2059 | -0.84 (-1.70, 0.01) | 0.0518 |
| Current users |  |  |  |  |  |  |  |  |
| 1 use/week | **-1.97 (-3.26, -0.68)** | **0.0028** | **2.82 (1.05, 4.59)** | **0.0018** | -0.29 (-1.51, 0.92) | 0.6346 | 0.04 (-1.65, 1.73) | 0.9635 |
| 2-3 uses/week | **-3.76 (-5.21, -2.32)** | **<.0001** | **3.53 (1.71, 5.35)** | **0.0001** | -0.53 (-1.89, 0.82) | 0.4379 | 0.30 (-1.44, 2.04) | 0.7362 |
| ≥ 4 uses/week | **-2.70 (-3.76, -1.64)** | **<.0001** | **3.72 (2.33, 5.10)** | **<.0001** | 0.10 (-0.89, 1.09) | 0.8444 | -0.53 (-1.85, 0.80) | 0.4344 |

<sup>\*</sup> Adjusted for age at interview, sex, and race/ethnicity,
<sup>†</sup> Model 1 + further adjustment for education level, poverty-to-income ratio, physical activity, serum cotinine, acculturation score, and NHANES cycles.

We found a significant interaction between sex and cannabis use in relation to PP (*P*=0.0878) and no interaction when modeling MAP (*P*=0.2084). Results from stratified analyses are shown in **Table 3**. PP was higher in males than in females, independent of cannabis use. We observed that cannabis use was significantly associated with PP in males, with a significant mean difference even in those with low frequency of use (i.e., 1 use/week). The mean difference between cannabis users and never users increased with the number of uses per week, being +4.5 mm Hg (*P*=0.0004) in those who reported 1 use/week, +4.9 mm Hg (*P*<0.0001) for 2-3 uses/week and +4.9 mm Hg (*P*<0.0001) for ≥ 4 uses/week. In females, only those who reported ≥ 4 uses/week showed a higher PP (+3.1 mm Hg; *P*=0.0050) compared with never users.

**Table 3:**
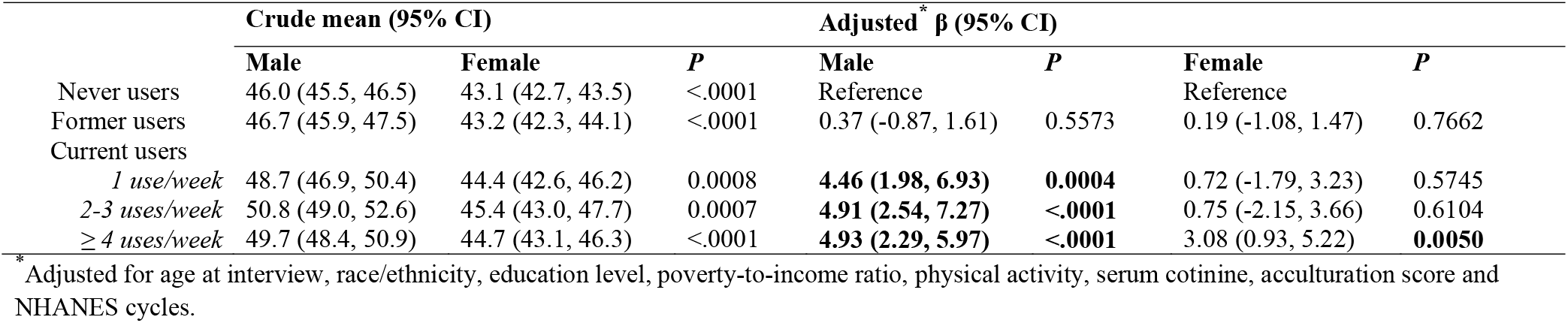
Sex-specific associations of cannabis use with pulse pressure in US adults, NHANES 2009-2018

| | Crude mean (95% CI) | | | Adjusted* $\beta$ (95% CI) | | | |
| --- | --- | --- | --- | --- | --- | --- | --- |
|  | Male | Female | *P* | Male | *P* | Female | *P* |
| Never users | 46.0 (45.5, 46.5) | 43.1 (42.7, 43.5) | <.0001 | Reference |  | Reference |  |
| Former users | 46.7 (45.9, 47.5) | 43.2 (42.3, 44.1) | <.0001 | 0.37 (-0.87, 1.61) | 0.5573 | 0.19 (-1.08, 1.47) | 0.7662 |
| Current users |  |  |  |  |  |  |  |
| *1 use/week* | 48.7 (46.9, 50.4) | 44.4 (42.6, 46.2) | 0.0008 | **4.46 (1.98, 6.93)** | **0.0004** | 0.72 (-1.79, 3.23) | 0.5745 |
| *2-3 uses/week* | 50.8 (49.0, 52.6) | 45.4 (43.0, 47.7) | 0.0007 | **4.91 (2.54, 7.27)** | **<.0001** | 0.75 (-2.15, 3.66) | 0.6104 |
| *≥ 4 uses/week* | 49.7 (48.4, 50.9) | 44.7 (43.1, 46.3) | <.0001 | **4.93 (2.29, 5.97)** | **<.0001** | 3.08 (0.93, 5.22) | **0.0050** |

\* Adjusted for age at interview, race/ethnicity, education level, poverty-to-income ratio, physical activity, serum cotinine, acculturation score and NHANES cycles.

## DISCUSSION

In this study that included US adults aged 18 to 59 years, we observed several key findings: first, compared with adults who never used cannabis, those who reported current use showed higher PP values and mean PP differences compared to never users increased with the frequency of cannabis use. Second, the positive association between cannabis use and PP remained in stratified analyses, but the impact of cannabis use on PP was more important in males than in females. Third, the association was statistically significant in males from low frequency of 1 use/week while a frequency of ≥ 4 use/week is required in females to observe significant difference with never users.

### Links with previous findings

No previous study in humans has investigated cannabis use in relation to PP and MAP. This makes it difficult to compare our results of a positive association between cannabis use and PP with those of previous reports. However, results from cross-sectional studies suggest a high prevalence of elevated BP in cannabis users, as compared with never users.^17^ Prospective studies suggested an inverse association between cannabis use and both systolic and diastolic BP.^20-22^ Our findings of positive association between cannabis use and PP suggest differential impact of cannabis exposure on systolic and diastolic BP. Such a differential impact is supported by findings from Jadoon et al. who suggest that CBD treatment in males reduces systolic BP without any impact on diastolic BP.^21^ In a cohort study including 11 men, Vandrey et al. found that BP increased significantly during periods of cannabis abstinence, the mean increase being more important in systolic BP (23 mm Hg) then in diastolic BP (12 mm Hg).

### Possible biological pathways (mechanisms) between cannabis use and widened PP

It is difficult to draw mechanistic conclusions from epidemiological studies such as the present one, based on heterogeneous populations self-administering a heterogeneous substance such as cannabis, in which the relative composition of the major pharmacologically active components, the cannabinoids, can change dramatically depending on the plant variety that is assumed. Furthermore, of the more than one hundred cannabinoids so far described to be present in the various cannabis varieties, the effect on cardiovascular endpoints in humans and mammals have been fully investigated only for THC (and its chemical artifact, CBN) and, more recently, CBD. Nevertheless, it is reasonable to assume that the recreational use of cannabis in more or less frequent users is due to the presence, in the self-administered substance, of predominant amounts of THC, possibly followed by CBD, since the presence of high amounts of CBD in cannabis preparations have been described to considerably reduce the psychotropic properties of THC.^36^ Both THC and CBD, via different mechanisms and receptors, are known to reduce MAP in both humans and laboratory animals,^37-39^ and hence the present finding of a positive correlation between frequency of cannabis use and higher PP, which should lead to higher MAP values, is counterintuitive. It is possible that chronic activation of CB1 by THC leads to its desensitization, and hence to higher MAP, a mechanism previously proposed also for the association between cannabis use and lower BMI and reduced incidence of type 2 diabetes despite the well-known positive effects of CB1 activation on energy balance.^40, 41^ However, to the best of our knowledge, no direct data is available to date on the effects of plant cannabinoids on PP, thus rendering difficult the interpretation of the present results. On the other hand, and possibly in agreement with the present findings, in women with depression, both diastolic and MAP were positively correlated with serum contents of endocannabinoids, anandamide and 2-arachidonoylglycerol, whilst there was no correlation between blood pressure and endocannabinoids in control subjects.^42^ Furthermore, depressed women had significantly higher systolic blood pressure than control subjects, suggesting that they may also have higher PP correlating with higher levels of endocannabinoids. Indeed, a possible confounding factor in the interpretation of our results may be represented by the fact that most of the mechanistic studies showing MAP lowering effects of THC, endocannabinoids, and other CB1 receptor activating agents, have been carried out in rodents with anxiety-induced high blood pressure, or in anaesthetized rats or in rats with hypertension.^43-46^

### Strengths and limitations of the current study

To our knowledge, this is the first study to investigate cannabis use in relation to PP and MAP. However, some study limitations need to be considered. First, the cross-sectional design of the study limits causal inference at a population level. The possibility that increased PP preceded the initiation of cannabis use cannot be ruled out. Second, cannabis use was self-reported in the NHANES study and exposure is then subject to misclassification bias. We cannot exclude such a misclassification to be sex-specific,^47^ leading to an appearance of effect modification by sex. Third, we missed data on the amount of cannabis smoked on each occasion. As a result, we could not approximate the true internal dose, and the direction of bias resulting from such misclassification cannot be predicted. Finally, no data on other recreational drugs have been available for adjustment, leading to potential residual confounding bias.

### Implication and perspectives

Recent data suggest that no safe cut-point for BP exists as organ damage may begin at prehypertension levels,^48, 49^ suggesting that slight differences in BP may result in long-term adverse effects. There is a dearth of longitudinal studies investigating the impact of cannabis use on PP. Previous cohort studies suffered from small sample size and sex imbalance. With evidence pointing to wide PP as a precursor of isolated systolic hypertension and early predictor of incident CVD,^2^ independent follow-up studies are needed to better understand the role of cannabis compounds on BP.

## CONCLUSION

In US adults aged 18 to 59 years, cannabis use is associated with widening of PP in males, and the mean difference in PP between cannabis users and never users increased with the frequency of use per week. In females, only those who reported ≥ 4 uses/week showed a higher PP compared with never users.

## Data Availability

Data are available on demand.

## Notes

### Competing Interest Statement

The authors have declared no competing interest.

### Funding Statement

No funding

### Author Declarations

NCHS Ethics Review Board (ERB) Approval

